# QuaID: Enabling Earlier Detection of Recently Emerged SARS-CoV-2 Variants of Concern in Wastewater

**DOI:** 10.1101/2021.09.08.21263279

**Authors:** Nicolae Sapoval, Yunxi Liu, Esther G. Lou, Loren Hopkins, Katherine B Ensor, Rebecca Schneider, Lauren B Stadler, Todd J Treangen

## Abstract

As clinical testing declines, wastewater monitoring can provide crucial surveillance on the emergence of SARS-CoV-2 variants of concern (VoC) in communities. Multiple recent studies support that wastewater-based SARS-CoV-2 detection of circulating VoC can precede clinical cases by up to two weeks. Furthermore, wastewater based epidemiology enables wide population-based screening and study of viral evolutionary dynamics. However, highly sensitive detection of emerging variants remains a complex task due to the pooled nature of environmental samples and genetic material degradation. In this paper we propose <u>quasi</u>-unique mutations for VoC <u>id</u>entification, implemented in a novel bioinformatics tool (QuaID) for VoC detection based on quasi-unique mutations. The benefits of QuaID are three-fold: (i) provides up to 3 week earlier VoC detection compared to existing approaches, (ii) enables more sensitive VoC detection, which is shown to be tolerant of >50% mutation drop-out, and (iii) leverages all mutational signatures, including insertions & deletions.

## Introduction

Wastewater monitoring is an invaluable tool for SARS-CoV-2 surveillance^1–8^. Despite multiple recent successes in VoC monitoring and detection from wastewater sequencing data^9–15^, there are multiple challenges associated with the nature of the environmental data. Since wastewater represents a pooled sample of multiple hosts, it harbors a diversity of SARS-CoV-2 variants that are currently circulating in the population^1,2,10,13^, including potentially previously unreported genotypes^16^. Furthermore, variant detection and phasing is further complicated by uneven genome coverage^2,17,18^ and environmental RNA degradation^5,19,20^ which render phased assembly extremely difficult^21–23^. Despite these challenges, detection of VoCs in wastewater samples is important for monitoring the emergence and spread of variants and informing public health response^4,5,11,24,25^. Current approaches for VoC detection in wastewater samples typically require sufficient depth and breadth of coverage of the variant genomes^9,12^, and therefore depend on a large fraction of the sample representing the variant genotype^10^, hampering early detection. Furthermore, most of the current approaches discard insertion and deletion (indel) information and only rely on single nucleotide variants (SNVs) associated with the VoC^9,12^. Finally, all approaches that rely on a database of previously collected SARS-CoV-2 genomes are biased by the contents of the database^26,27^, which can lead to both false negative and false positive calls at the inference stage^28^. This issue can be further amplified when the underlying database is not scrutinized for potential metadata errors^29,30^.

To address these issues, we developed QuaID: a computational pipeline for analyzing SARS-CoV-2 wastewater sequencing data and inferring presence of VoCs. We use empirical results on real Houston wastewater data and simulated data to validate the efficacy of our software, and compare it to Freyja^9^, another state-of-the-art tool for VoC detection. Our key goal is achieving sensitivity to newly emerging variants in scenarios where coverage breadth and depth can be uneven, and the VoC-associated genomes are present at low abundances. We also leverage the indel data that can be inferred from the multiple sequence alignments to further improve the robustness of our VoC detection approach.

## Results

Between February 23, 2021 and May 5th, 2022 we collected, processed, and analyzed 2,637 wastewater samples from the fifth-most populous metropolitan area in the US: Houston, Texas. Samples were collected weekly from 39 wastewater treatment plants (WWTPs, Supplementary Table 1, Supplementary Figure 1) distributed throughout the city of Houston and servicing more than 2 million Houston residents^18^. During the study period, the VoC detection signal clearly reflected the three major variants that affected Houston - Alpha, Delta, and Omicron (Figure 1A). QuaID was able to detect the Delta VoC two weeks prior to the first sequenced clinical sample in Texas (marked by star in Figure 1B) and continued to provide detection signal for the four subsequent weeks after the first sequenced clinical sample (2021-04-05 to 2021-05-03).

**Figure 1.**
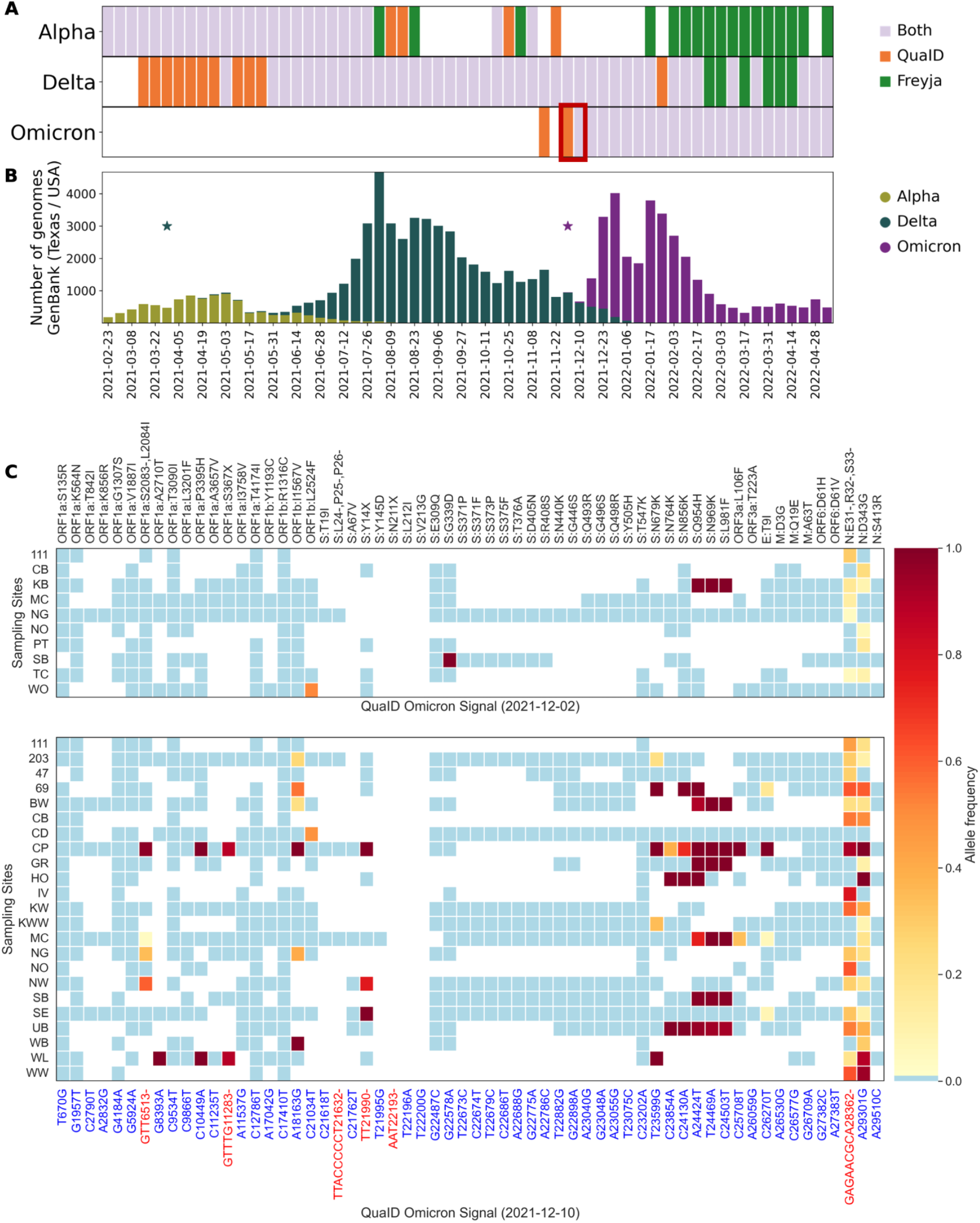
Detection of Alpha, Delta, and Omicron VoCs in Houston, TX wastewater. **A**. Early detection of the emerging variants of concern in Houston wastewater provided by QuaID and Freya pipelines. For Omicron and Delta variants, QuaID provided earlier detection. Each week is presented as the aggregate signal from the 39 WWTPs with detections being reported if at least 2 WWTPs had any QuaID signal or had any non-zero abundance of the VoCs reported by Freyja. **B**. Variant prevalence in the clinical data over the study period obtained from GenBank and restricted to Texas. Stars indicate the first occurrences of a Delta variant genome (yellow) and an Omicron variant genome (red). **C**. Heatmaps of WWTPs with detected Omicron variant quasi-unique mutations the week of December 2nd, 2021 (top) and December 10th, 2021 (bottom) in Houston. Blanks indicate lack of sequencing data, blue color indicates no mutation detected, and the gradient shows the allele frequency for detected mutations.

In contrast, Freyja reliably picked up the Delta signal only once the VoC became more prevalent. Similarly for the Omicron VoC, QuaID detected the presence of the variant in wastewater two weeks prior to the first clinical sample collection date, while Freyja required an additional week after the first clinical sample to detect Omicron presence.

We further investigated the early detection of the Omicron variant in Houston wastewater by visualizing a heatmap of variant calls (Figure 1C) and examining the multiple sequence alignment (MSA) of SARS-CoV-2 Omicron variant genomes available on GISAID^31^ in early December 2021. We observed 50% (5 out of 10) of the samples with Omicron presence for the week of December 2nd, 2021 contained the 9bp deletion (N:DEL31/33), which is a stable mutation (95.1% prevalence among all Omicron genomes^32^) for the Omicron variant (Figure 1C). Since the current version of Freyja relies on the UShER^33^ phylogenetic tree for its designation of mutational signatures, no deletions are used in the inference process, highlighting one of the reasons for the delayed detection of the Omicron variant. In the subsequent week, December 10th, 2021, when both Freyja and QuaID reported the presence of the Omicron variant in the wastewater, the N:DEL31/33 mutation was present in 16 of 23 sites with detections (Figure 1C), and for one of the samples with no deletion there was no coverage in the region flanking the deletion (Figure 1C, Sampling Site SB: Sims Bayou North).

To further examine sensitivity of the QuaID and Freyja to degradation of the sequencing data, we constructed a simulated data experiment in which we varied the fraction of SNVs dropped out from the variant calling results. In the real wastewater sequencing data, 37.7% of all samples had less than 25% of the SNVs associated with the Omicron VoC via UShER barcodes covered by at least one read (Supplementary Figure 2B), and 24.4% of samples had less than 10% of all Omicron-associated SNVs with at least one read. Thus, we constructed three simulation scenarios with each retaining 10%, 25%, or 50% of all SNV calls at random. Our results show that due to the inclusion of deletion information in the inference process, QuaID remained sensitive even when only 10% of all SNV calls were retained, while Freyja required at least 50% of the calls to be included to reliably detect the VoC presence. In particular, when only 10% of all SNV calls were retained, QuaID still detected the presence of Delta and Omicron VoCs reliably, and Alpha and Gamma VoCs sparsely, while Freyja failed to estimate the abundance of any of the VoCs (Alpha, Delta, Gamma, and Omicron) present in the simulated samples (Figure 2A, Supplementary Figures 3A-6A). Furthermore, when 25% of all SNVs were retained, QuaID identified the present VoCs in the majority of the simulated samples, while Freyja provided sparse detection in the samples dominated by a single VoC (Figure 2B). Finally, when 50% of all SNVs are retained, Freyja detected most of the VoCs present in the samples, and in several instances recovered the correct relative abundance. However, even in this scenario 8 Omicron dominated samples failed to be correctly identified by Freyja, while QuaID correctly inferred the presence of the VoC. Additionally, we observe that the stability of the coverage for the N:DEL31/33 is further empirically supported by our data, which indicated that among all samples more than 61% have at least 10 reads that cover the bases immediately flanking the deletion (Supplementary Figure 2C).

**Figure 2.**
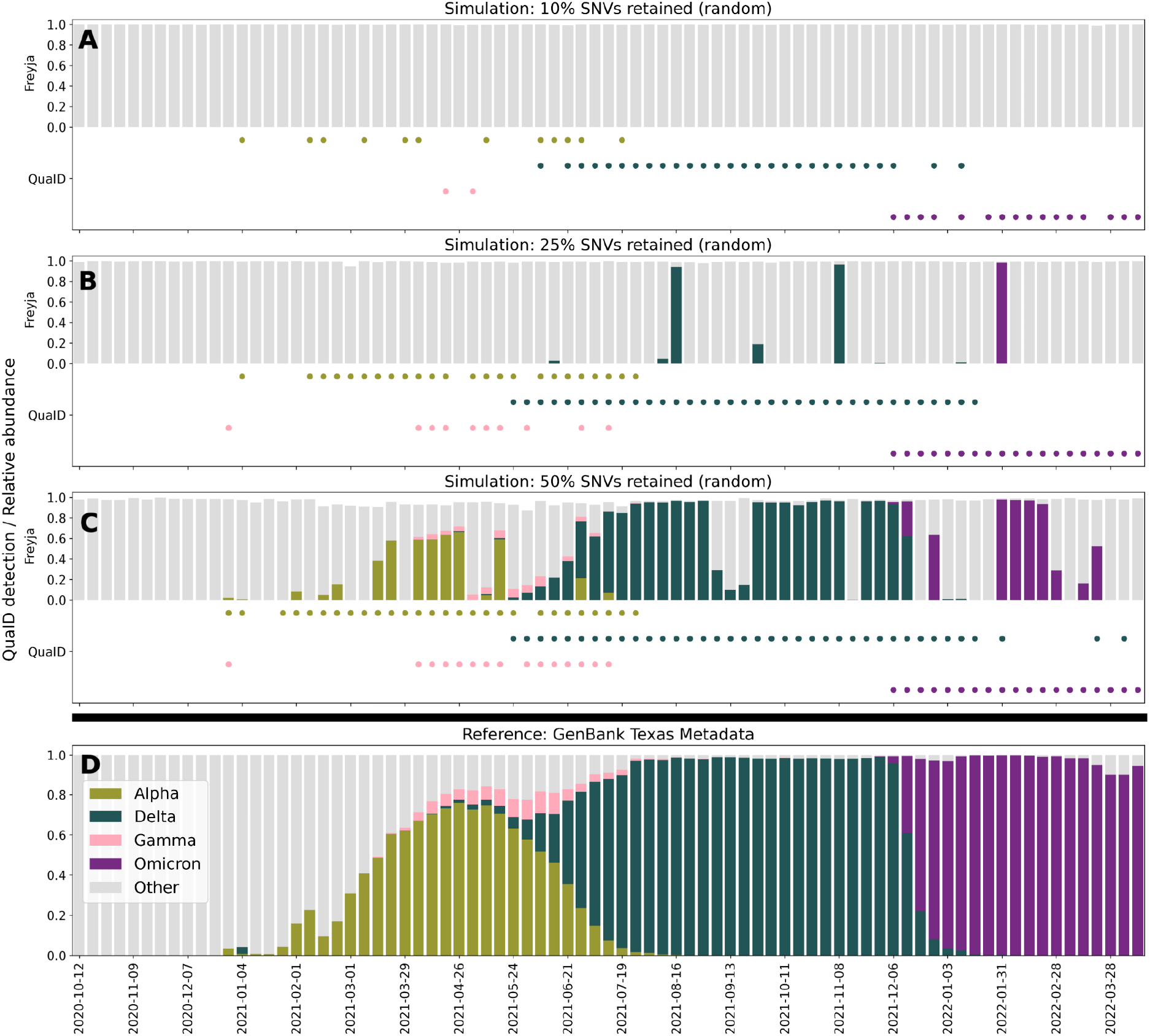
Detection of VoCs in simulated data at various levels of SNV dropout. **A**. Freyja relative abundance estimates and QuaID detection signal on simulated data from GenBank (USA/TX) with 10% of all SNVs retained at random. Freyja is unable to detect any of the four (Alpha, Delta, Gamma, Omicron) VoCs. **B**. Freyja relative abundance estimates and QuaID detection signal on simulated data from GenBank (USA/TX) with 25% of all SNVs retained at random. Freyja sparsely detects major VoCs (Delta, Omicron). QuaID detections are less sparse for all VoCs. **C**. Freyja relative abundance estimates and QuaID detection signal on simulated data from GenBank (USA/TX) with 50% of all SNVs retained at random. **D**. Metadata from GenBank (USA/TX) showing the fraction of genomes belonging to different VoCs for any given week. In this simulated experiment the fractions shown correspond to true relative abundances in the simulated mixture.

## Discussion

Wastewater monitoring of the SARS-CoV-2 variant emergence and spread offers unique benefits based on the early detection of the variant arrival prior to the clinical data^3,34,35,15,25^, and broad surveillance coverage of the population^20,34,36,37^. QuaID offers a highly sensitive pipeline for VoC detection using wastewater SARS-CoV-2 sequencing data. In comparison to one of the leading tools for analyzing SARS-CoV-2 wastewater sequencing data for VoC detection, QuaID demonstrated superior sensitivity. This is particularly important given that the underlying sample quality varies and the depth and breadth of coverage of amplicon sequencing data can vary widely across samples^18^. Furthermore, the ability to leverage indel information in the inference process makes QuaID overall more robust than approaches that rely solely on SNVs^9,15^. Methods that currently rely on phylogenetic placement or utilize phylogenetic trees inferred by tools like UShER^33^ will likely continue to lack support for indel based detection.

While full phylogenetic placement remains the gold standard for strain level analyses of mixed population samples^38^, challenges posed by the data quality of SARS-CoV-2 wastewater samples limit applicability of such approaches. Thus, QuaID is designed as an early detection tool that does not perform full phylogenetic placement of reads. Additionally, since our main goal was high sensitivity to emerging variants in scenarios where the underlying mutational signal is low, QuaID treats each observed mutation as an independent event, and hence is not in its current form suited to perform relative abundance estimates.

Since QuaID relies on the definition of the quasi-unique mutations for the VoCs, the selection of thresholds for mutation inclusion into the quasi-unique set for a given lineage and for the exclusion of a mutation from this list if found in another lineage affects the sensitivity and specificity of our method. Namely, setting a higher inclusion threshold (i.e. requiring that more of the target lineage genomes contain the mutation) and lower exclusion threshold (i.e. allowing a lower mutational prevalence in other lineages to exclude mutation from the list) will increase specificity, but decrease sensitivity of the method. Our choice of the 50% as the threshold for both inclusion and exclusion is motivated by the prior work for bacterial identification in the field of metagenomics^39,40^. Furthermore, given that the current genomic database contains more than 10 million SARS-CoV-2 sequences with major variants contributing millions of sequences, setting restrictive thresholds (e.g. requiring that all genomes classified as a given lineage contain the mutation, i.e. setting inclusion threshold to 100%; or alternatively requiring that no genomes outside the target lineage contain the mutation) can result in empty quasi-unique sets for certain lineages.

We envision QuaID to be one of several tools routinely employed in wastewater monitoring efforts. For example, QuaID could be used in parallel with Freyja to achieve high sensitivity for detecting emerging variants, and relative abundance estimates of the dominant circulating variants. Furthemore, future work on extending the framework of QuaID and other tools to other pathogens that can be detected in the wastewater can enable sensitive and continuous environmental monitoring^41,42^ beyond the COVID-19 pandemic. Finally, given the multitude of technical challenges posed by the inherent variability and quality of wastewater sequencing data, we believe that establishing extensive sets of simulated and synthetic datasets that emulate challenges in variant calling in wastewater samples are required to further expand our understanding of how RNA degradation, sample preparation and storage techniques, and sequencing protocols affect the downstream data and analyses.

## Methods

### Wastewater sample collection, RNA extraction, and sequencing

Houston Water collected and provided weekly 24-hour time-weighted composite influent (raw wastewater) samples from 39 wastewater treatment plants (WWTPs) in Houston covering a service area of approximately 580 miles^2^ and serving over 2.3 million people. In total, 2,637 samples were analyzed. SARS-CoV-2 was concentrated in wastewater samples using an electronegative filtration method as previously described^43^. We followed the same RNA extraction, library preparation, and sequencing protocols, as described in prior work^18^. Details on WWTP sample sites, and methods regarding sample collection procedures, and quantification of SARS-CoV-2 in wastewater samples can also be found in our previous publication^18^. Estimates of the viral load are provided by the Houston Health Department following the same methodology as outlined in the SARS-CoV-2 Wastewater Monitoring Dashboard^44^. Raw sequencing reads for the samples used in this study can be found in SRA under BioProject accession: PRJNA796340.

### Amplicon sequencing data processing

We processed the MiSeq paired-end data through a standard sequence of steps consisting of quality control report generation (FastQC^45^, default parameters), quality and adapter trimming (BBDuk^46^, quality trimming both ends of the read with threshold 15, and trimming standard PhiX adapter sequences), read mapping (BWA MEM^47^, default parameters), and primer site soft clipping (iVar^48^, ARTIC v3^49^ primer scheme, minimum quality threshold 15). The summary overview of the whole processing pipeline is presented in Figure 3.

**Figure 3.**
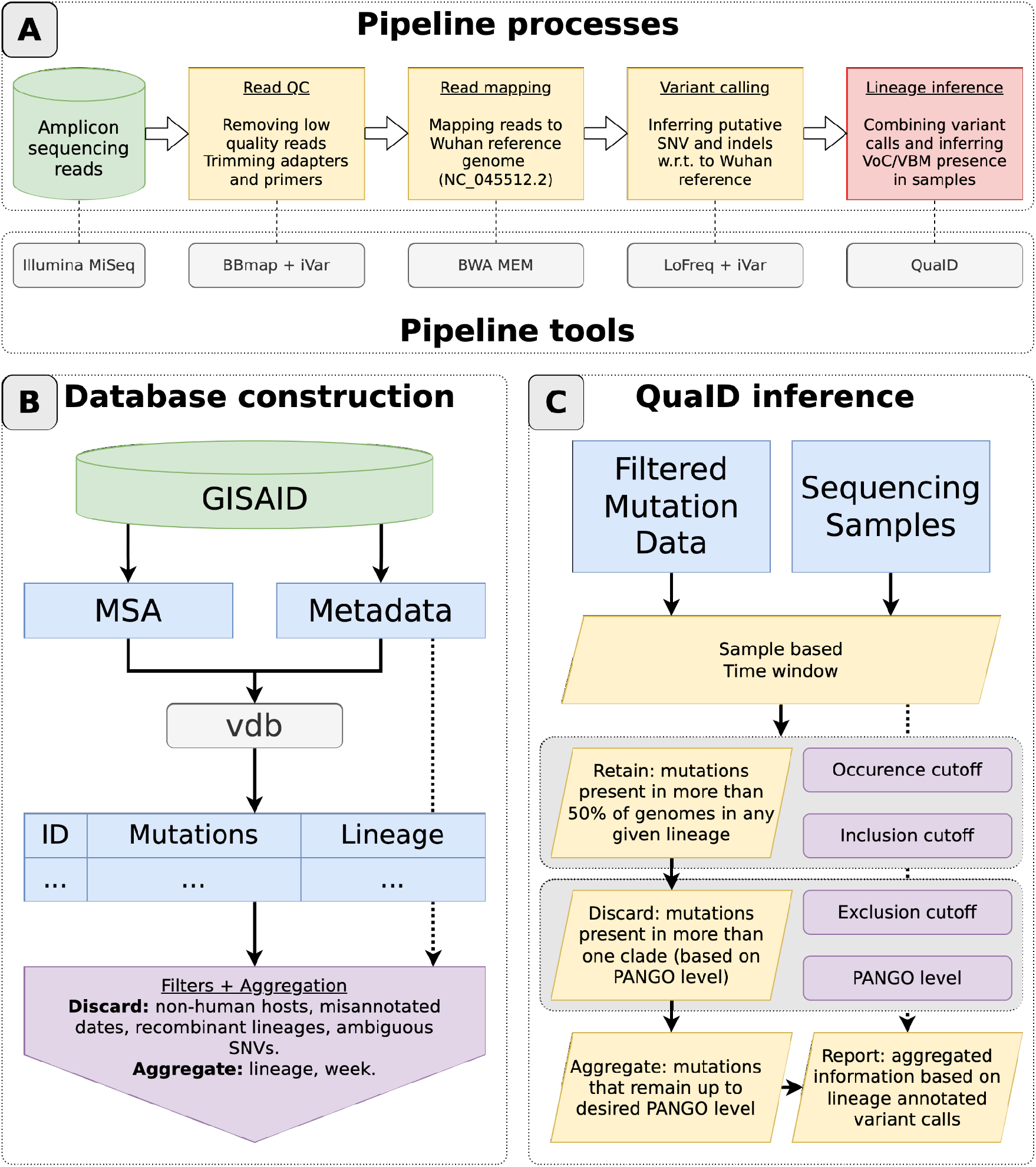
Overview of QuaID computational approach. **A**. Overview of the complete sequencing data processing pipeline employed in the analysis of the Houston wastewater SARS-CoV-2 sequencing data. **B**. Schematics overview of the mutation database construction and sanitation used by QuaID. MSA and metadata obtained from GISAID are first pre-processed with vdb to extract mutations, and then custom Python code is used to perform additional filtering and aggregation procedures. **C**. QuaID VoC/VBM inference process overview. Parameters that affect described subroutines (yellow parallelograms) are provided in the purple rounded rectangles.

### Variant calling

We obtained two sets of variant calls for each sample: one with iVar^48^ (minimum quality 20, minimum allele frequency 0) and the other with LoFreq^50^ (after adjusting quality scores for indel calling with the ‘lofreq indelqual --dindel’ call, variant calling parameters are set to default). Both variant callers were configured to output all variant calls regardless of the allele frequency. We then used custom Python code to perform a variant call merge-and-filter operation which retained only those variant calls that were supported by both variant callers and had an allele frequency equal or above the user-defined threshold (default: 0.02) according to at least one of the two variant callers (while allele frequency estimates are typically close between the two variant callers differences of <0.01 occur).

### Sequence database sanitation

Prior to the subsequent analysis we used metadata obtained from GISAID website to filter out sequences that were marked as incomplete or that had an associated host other than *Homo Sapiens*. Additionally, VoCs with a large amount of clinical sequencing data available (Alpha, Delta, Omicron) are more prone to human error in the metadata entries. Thus, we implemented a filter that removed any genomes: annotated as Alpha with submission date prior to September 3rd, 2020, annotated as Delta with submission date prior to March 1st, 2021, and annotated as Omicron with submission date prior to September 1st, 2021 (first detection dates based on <u>cov-lineages.org</u> VoC reports). Finally, we excluded all recombinant PANGO lineages^51^ (X*) from the analysis.

### Mutation database construction

We used the pre-generated MSA file from GISAID to extract all mutations using vdb^52^ in nucleotide mode with ambiguous bases included. We then trimmed the resulting list of mutations using the vdb trim command. Finally, we linked the resulting mutation list with the metadata based on the genome accession IDs, and the resulting data were aggregated by week and lineage through custom Python code. Additionally, any SNVs that resulted in an ambiguous base call (e.g. N, W, S, etc.) were removed from the database (summary view provided in Supplementary FIgure 2B). The resulting data were used as the mutation tables to calculate prevalence of mutations in PANGO lineages^51^ over a user-defined time window (default: 4 weeks).

### Quasi-unique mutations

For each lineage and mutation combination, the prevalence of the mutation occurring in the corresponding lineage’s genomes was calculated and then converted to a fraction of all genomes assigned to the lineage. Mutations that appeared in more than 50% of all genomes for a single lineage (i.e. not appearing in any other lineage at 50% or more) were considered quasi-unique for that lineage. The above choice of inclusion (what fraction of genomes in the lineage must have the mutations) and exclusion (what fraction of genomes in any other lineage precludes the mutation from being selected) corresponds to the definition of a consensus genome but can be modified to arbitrary values by the end user. Setting stricter thresholds (requiring more of the target lineage genomes to have a mutation) will lead to smaller sets of quasi-unique mutations of high confidence, trading of sensitivity for specificity. Furthermore, since often we want to report detections at a higher level (e.g. any Omicron sub-lineage as opposed to a specific leaf node like BA.2.1) when determining which genomes are used for the exclusion rule, all the genomes that come from the same sub-clade at a fixed level (default: 4) in PANGO hierarchy are omitted from the exclusion check. Thus, mutations common to BA.1 and BA.2 can still be considered as quasi-unique for the Omicron VoC. Note that since vdb reports out deletions and we only filter out ambiguous SNVs, a quasi-unique mutation can be a deletion. Additionally, in order to reduce potential noise from extremely rare lineages, we omit any lineages which have less than a user-defined count of genomes (default: 2) within the designated time window. An overview of these processes is presented in Figure 3C. Finally, for each quasi-unique mutation QuaID estimated its predictive power as the posterior probability of observing a particular lineage given the observed mutation. Formally, for a lineage of interest *l* and the quasi-unique mutation *m* we computed *P*(*l*|*m*)using Bayes’ theorem 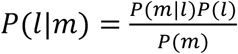. We let *P*(*m*) to be the ratio of the number of genomes with the mutation *m* observed to the total number of genomes in the database. Next, we let *P*(*l*) to be the ratio of the number genomes belonging to the lineage *l* and the total number of genomes. Finally, we let *P*(*m*|*l*) to be the fraction of genomes in the lineage *l* containing the mutation *m*. While we did not provide any filtering based on the estimated predictive power of the quasi-unique mutations, these probabilities can be used in the downstream analyses to improve the interpretations of the detection signal provided by QuaID.

### Mutational signature aggregation

Since the PANGO lineage hierarchy continuously expands potentially introducing new sub-levels for any lineage, it is useful to aggregate quasi-unique mutations into sets that correspond to a node at a fixed level of the hierarchy. For example, Omicron variant is defined as any descendant of B.1.1.529 PANGO lineage, and thus Omicron corresponds to level 4 in the hierarchy. When aggregating quasi-unique mutational signatures up to a given level, we took the union of all descendant lineage quasi-unique mutation sets. Note that the aggregation step always uses the same level of the hierarchy as the exclusion step of the quasi-unique mutation set construction.

### Variant of concern detection

Given a wastewater-based sequencing sample collected on a given date *D*, we constructed the corresponding sets of quasi-unique mutations in the time-window prior to and including weeks up to date *D* (in case when there is no database information for the week(s) immediately preceding the target date *D*, the last available time-window was used). Then we merged the filtered set of variant calls for the sample with the quasi-unique set of mutations with the key set to the nucleotide change. We also filtered out any SNVs from the sample that result in synonymous mutations. Once the combined data is obtained, we report for each sample the total combined allele frequency and total count of observed quasi-unique mutations, as well as the total possible number of quasi-unique mutations for the variants of interest at the desired level. Additionally, we reported what percentage of the quasi-unique mutation sites had coverage (with deletions being evaluated based on the genomic positions flanking the deletion) in order to distinguish between the “no detection” and “no coverage” scenarios.

## Data Availability

Data is available upon request.
Sequencing data available online on NCBI SRA Bioproject PRJNA796340.

## Data availability

Raw sequencing data used in this study is available on SRA under BioProject accession PRJNA796340. Software developed in this manuscript and used to generate the results is available at https://gitlab.com/treangenlab/quaid.

## Acknowledgements

The authors thank the GISAID contributors who provided the SARS-CoV-2 assemblies. We thank Roger Sealy, Pamela Brown, Ryker Penn, and Yanlai Lai (Houston Health Department) for their assistance in sample collection and sequencing. The authors also would like to thank Lauren Bauhs, Russell Carlson-Stadler, Madeline Wolken, Kyle Palmer, Whitney Rich (Rice University) for their assistance in sample collection, processing, and analysis. This work was supported by the Houston Health Department. Funding sources for sequencing by the Houston Health Department were CDC ELC Enhanced Detection, CDC ELC Enhanced Detection Expansion, and CDC ELC Advanced Molecular Detection. N.S., Y.L. and T.J.T. were supported in part by the C3.ai DTI, Centers for Disease Control (CDC) contract 75D30121C11180 and P01-AI152999 NIH award. E.G.L. and L.B.S. were supported in part by the National Science Foundation (CBET 2029025), and seed funds from Rice University. K.B.E. was supported in part by National Institute of Environmental Health Sciences, R01ES028819.

## Author contributions

N.S. has conducted data analyses, designed and implemented software methods, designed and implemented the experiments. Y.L. has tested and implemented software methods. Y.L., E.G.L., and R.S. have conducted data collection and analyses. L.H., K.B.E., L.B.S., T.J.T. have overseen the data collection and analyses, designed and reviewed the experiments. All authors have participated in writing and reviewing the manuscript.

