## Supplementary materials for "QuaID: Enabling Earlier Detection of Recently Emerged SARS-CoV-2 Variants of Concern in Wastewater"

**Supplementary Table 1.** Wastewater treatment plants sampled, abbreviations, average flow rates, service populations, and geographic service areas

| Wastewater treatment plant | Abbreviation | Flowrate, MGD (AVG ± SD) | Population | Average gal/cap/day | Area, square miles |
|---|---|---|---|---|---|
| 69th Street | 69 | 80.03 ± 21.77 | 551,150 | 145 | 96.72 |
| Almeda Sims | AS | 13.76 ± 11.93 | 117,968 | 117 | 54.93 |
| Beltway | BW | 6.93 ± 4.70 | 70,900 | 98 | 9.76 |
| Cedar Bayou | CD | 0.78 ± 0.42 | 1,722 | 453 | 3.27 |
| Chocolate Bayou | CB | 4.03 ± 4.19 | 37,359 | 108 | 14.61 |
| Clinton Park | CP | 0.69 ± 0.81 | 3,825 | 180 | 4.14 |
| Easthaven | EH | 1.89 ± 1.85 | 16,030 | 118 | 4.78 |
| FWSD#23 | 23 | 3.09 ± 2.52 | 40,689 | 76 | 15.14 |
| Forest Cove | FC | 0.28 ± 0.13 | 4,170 | 67 | 2.73 |
| Greenridge | GR | 3.06 ± 3.06 | 28,742 | 106 | 6.87 |
| Homestead | HO | 1.58 ± 1.53 | 9,375 | 169 | 6.12 |
| Imperial Valley | IV | 1.75 ± 0.70 | 16,804 | 104 | 2.22 |
| Intercontinental Airport | IA | 1.91 ± 0.63 | 2,408 | 793 | 38.73 |
| Keegans Bayou | KB | 14.25 ± 10.31 | 124,000 | 115 | 13.78 |
| Kingwood Central | KW | 3.49 ± 1.46 | 52,055 | 67 | 23.04 |
| Kingwood West | MG | 0.61 ± 0.20 | 2,589 | 236 | 2.6 |
| MUD#203 | 203 | 0.38 ± 0.12 | 4,010 | 95 | 2.57 |
| Metro Central | MC | 1.99 ± 1.64 | 20,161 | 99 | 9.86 |
| Northbelt | NO | 2.37 ± 1.49 | 12,892 | 184 | 15.79 |
| Northeast | NE | 3.88 ± 4.25 | 33,102 | 117 | 14.41 |
| Northgate | NG | 2.75 ± 1.03 | 19,867 | 138 | 3.6 |
| Northwest | NW | 9.99 ± 5.84 | 95,600 | 104 | 22.62 |
| Park Ten | PT | 0.62 ± 0.31 | 5,497 | 113 | 2.19 |
| Sagemont | SG | 4.52 ± 3.49 | 20,608 | 219 | 5.9 |
| Sims Bayou South* | SS | 23.93 ± 18.22 | 109,414 | 219 | 47.84 |
| Sims Bayou North* | SB | 8.22 ± 7.54 | 109,414 | 75 | 47.84 |
| Southeast | SE | 4.88 ± 4.85 | 32,485 | 150 | 9.06 |
| Southwest | SW | 37.59 ± 26.39 | 293,227 | 128 | 38.72 |
| Tidwell Timbers | TT | 0.11 ± 0.06 | 1,133 | 97 | 0.57 |

| | | | | | |
|---|---|---|---|---|---|
| Turkey Creek | TC | 7.00 ± 4.85 | 59,188 | 118 | 10.46 |
| Upper Brays | UB | 10.33 ± 7.44 | 97,918 | 105 | 12.81 |
| WCID#111 | 111 | 2.24 ± 0.28 | 20,920 | 107 | 3.35 |
| WCID#47 | 47 | 3.36 ± 2.28 | 33,645 | 100 | 6.27 |
| WCID#76 | 76 | 0.37 ± 0.22 | 976 | 379 | 0.5 |
| West District | WD | 10.06 ± 6.62 | 85,129 | 118 | 17.86 |
| West Lake | WL | 0.20 ± 0.07 | 600 | 333 | 0.53 |
| Westway | WW | 0.40 ± 0.18 | 3,623 | 110 | 0.99 |
| White Oak | WO | 1.84 ± 0.91 | 20,758 | 89 | 3.31 |
| Willowbrook | WB | 1.28 ± 0.52 | 8,610 | 149 | 3.01 |
| **TOTAL** | | **272.23 ± 159.04** | **2,168,563** | **162 ± 133 (AVG ± STDEV)** | **532** |

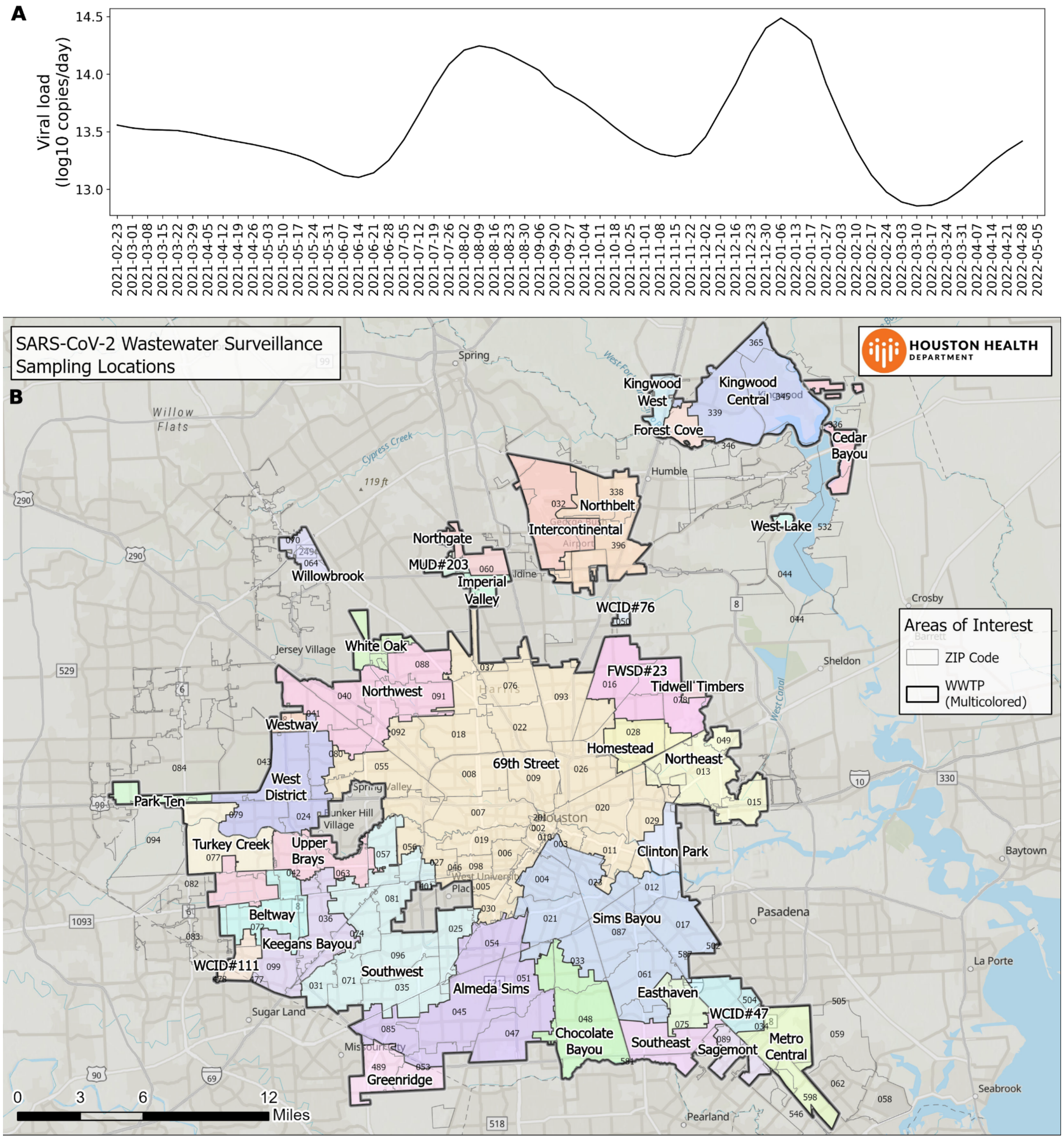


**Supplementary Figure 1. A.** Estimated viral load in Houston wastewater in log-scale of viral copies per day. Trend in viral load matches up with the variant associated waves of infection. **B.** Map of the sewersheds and wastewater treatment plants in the city of Houston.

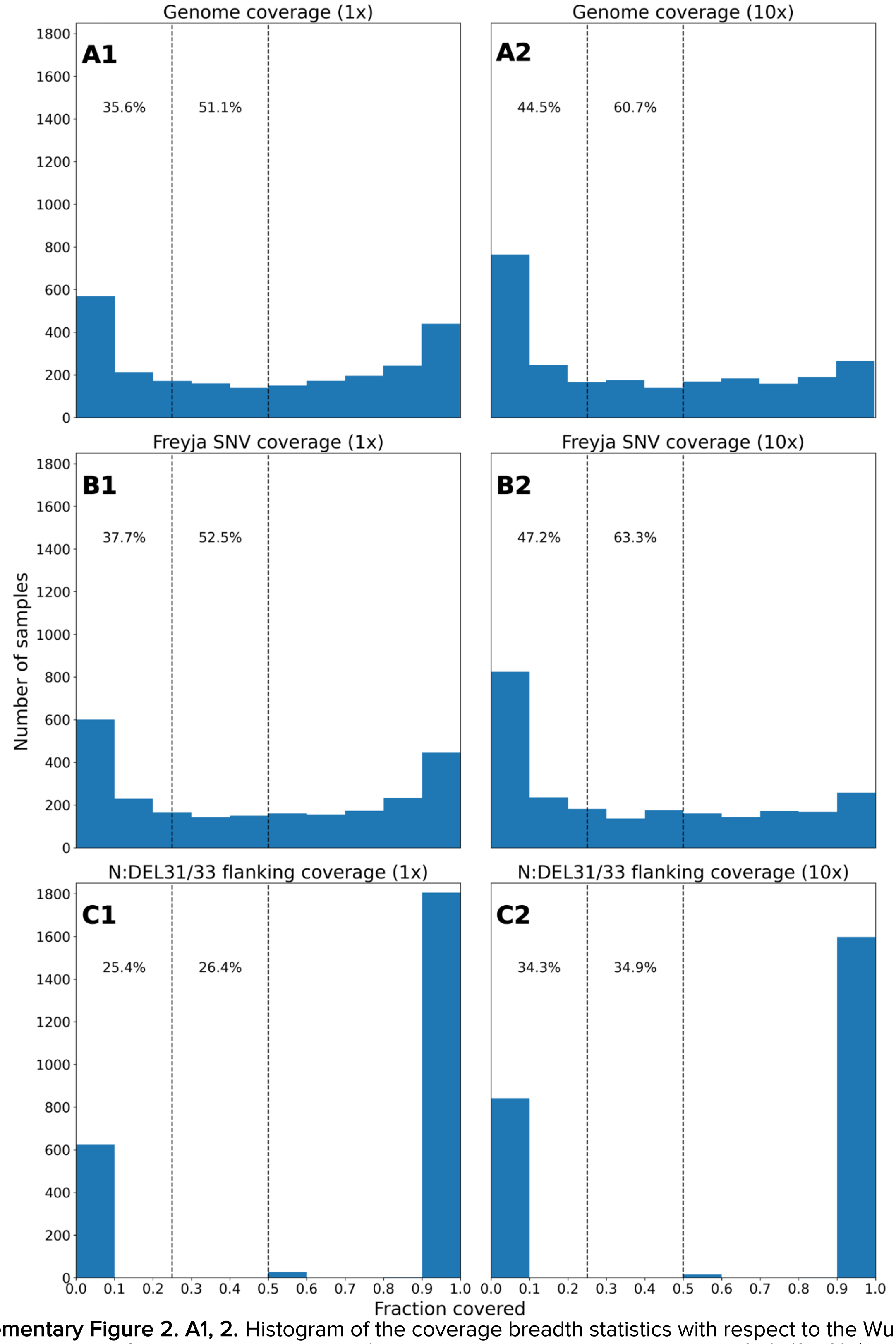


**Supplementary Figure 2. A1, 2.** Histogram of the coverage breadth statistics with respect to the Wuhan reference genome. Cumulative percentage of samples with coverage breadth up to 25% (35.6%/44.5% of all samples) and 50% (51.1%/60.7% of all samples) is noted above the histogram. **B1, 2.** Histogram of the fraction of the SNVs used by Freyja to detect Omicron VoC that have coverage. Cumulative percentage of samples with coverage fraction up to 25% (37.7%/52.5% of all samples) and 50% (47.2%/63.3% of all samples) is noted above the histogram. **C1, 2.** Histogram of the fraction of the flanking positions (6 in total) for the N:DEL31/33 that have coverage. Cumulative percentage of samples with coverage fraction up to 25% (25.4%/26.4% of all samples) and 50% (34.3%/34.9% of all samples) is noted above the histogram.

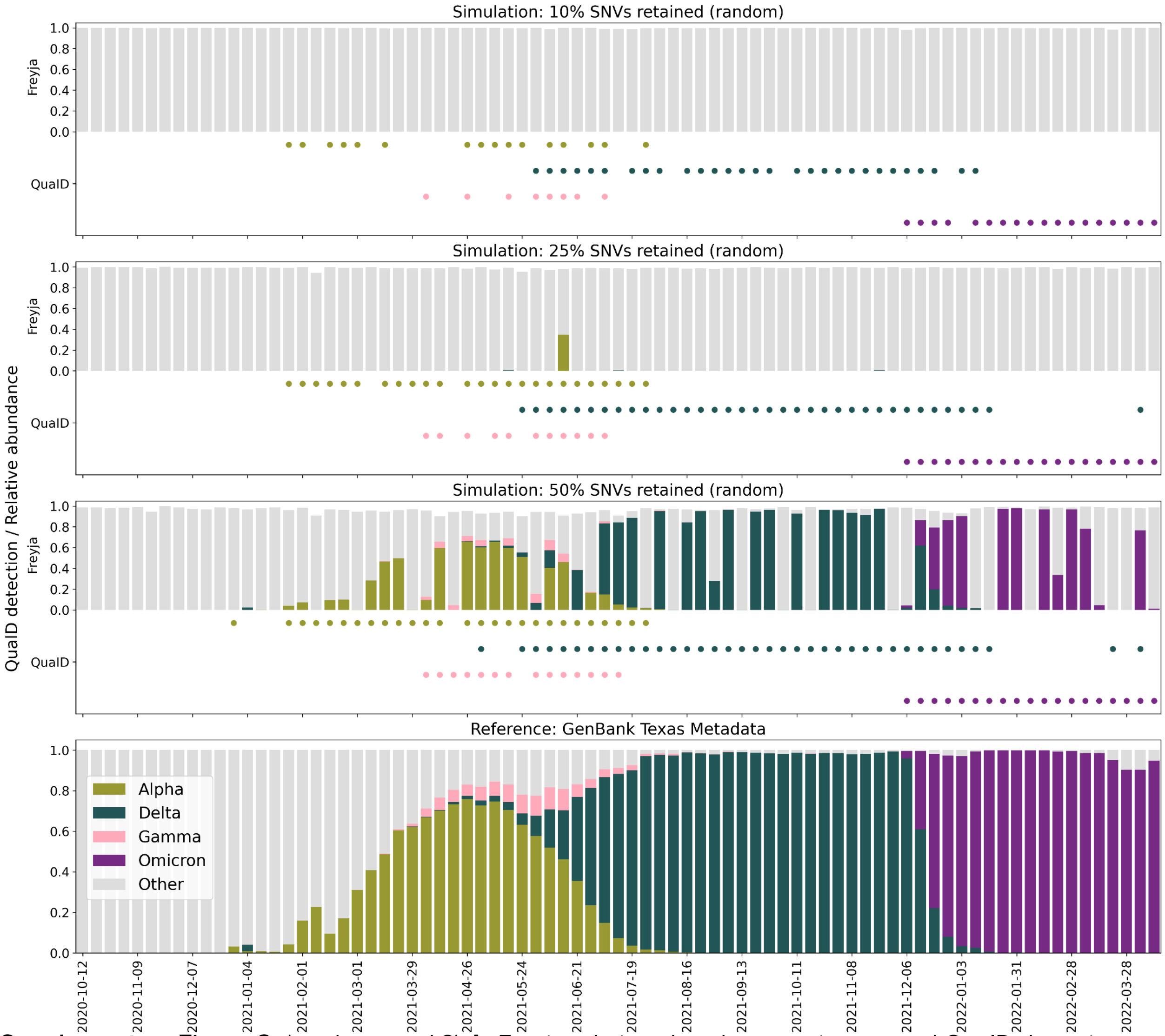


**Supplementary Figure 3.** (random seed 3) **A.** Freyja relative abundance estimates and QualD detection signal on simulated data from GenBank (USA/TX) with 10% of all SNVs retained at random. Freyja is unable to detect any of the four (Alpha, Delta, Gamma, Omicron) VoCs. QualD detection is sparse, in particular for the Gamma variant. **B.** Freyja relative abundance estimates and QualD detection signal on simulated data from GenBank (USA/TX) with 25% of all SNVs retained at random. Freyja sparsely detects major VoCs (Delta, Omicron). QualD detections become less sparse for all VoCs. **C.** Freyja relative abundance estimates and QualD detection signal on simulated data from GenBank (USA/TX) with 50% of all SNVs retained at random. Freyja detections become dense, and in some cases abundance estimates correctly reflect simulated abundance profiles. QualD remains highly sensitive with respect to early detection. **D.** Metadata from GenBank (USA/TX) showing the fraction of genomes belonging to different VoCs for any given week. In this simulated experiment the fractions shown correspond to true relative abundances in the simulated mixture.

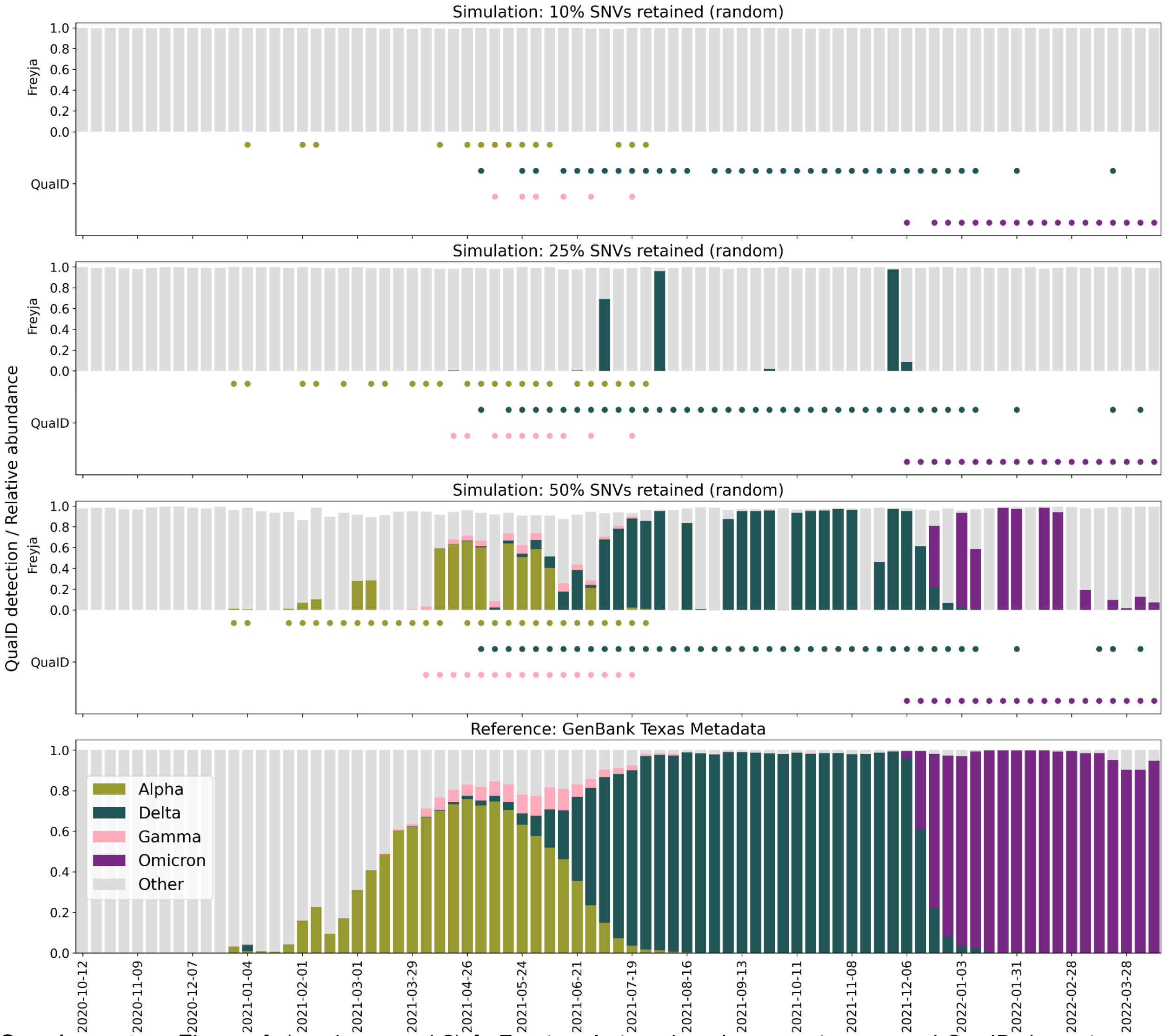


**Supplementary Figure 4.** (random seed 8) **A.** Freyja relative abundance estimates and QualD detection signal on simulated data from GenBank (USA/TX) with 10% of all SNVs retained at random. Freyja is unable to detect any of the four (Alpha, Delta, Gamma, Omicron) VoCs. QualD detection is sparse, in particular for the Gamma variant. **B.** Freyja relative abundance estimates and QualD detection signal on simulated data from GenBank (USA/TX) with 25% of all SNVs retained at random. Freyja sparsely detects major VoCs (Delta, Omicron). QualD detections become less sparse for all VoCs. **C.** Freyja relative abundance estimates and QualD detection signal on simulated data from GenBank (USA/TX) with 50% of all SNVs retained at random. Freyja detections become dense, and in some cases abundance estimates correctly reflect simulated abundance profiles. QualD remains highly sensitive with respect to early detection. **D.** Metadata from GenBank (USA/TX) showing the fraction of genomes belonging to different VoCs for any given week. In this simulated experiment the fractions shown correspond to true relative abundances in the simulated mixture.

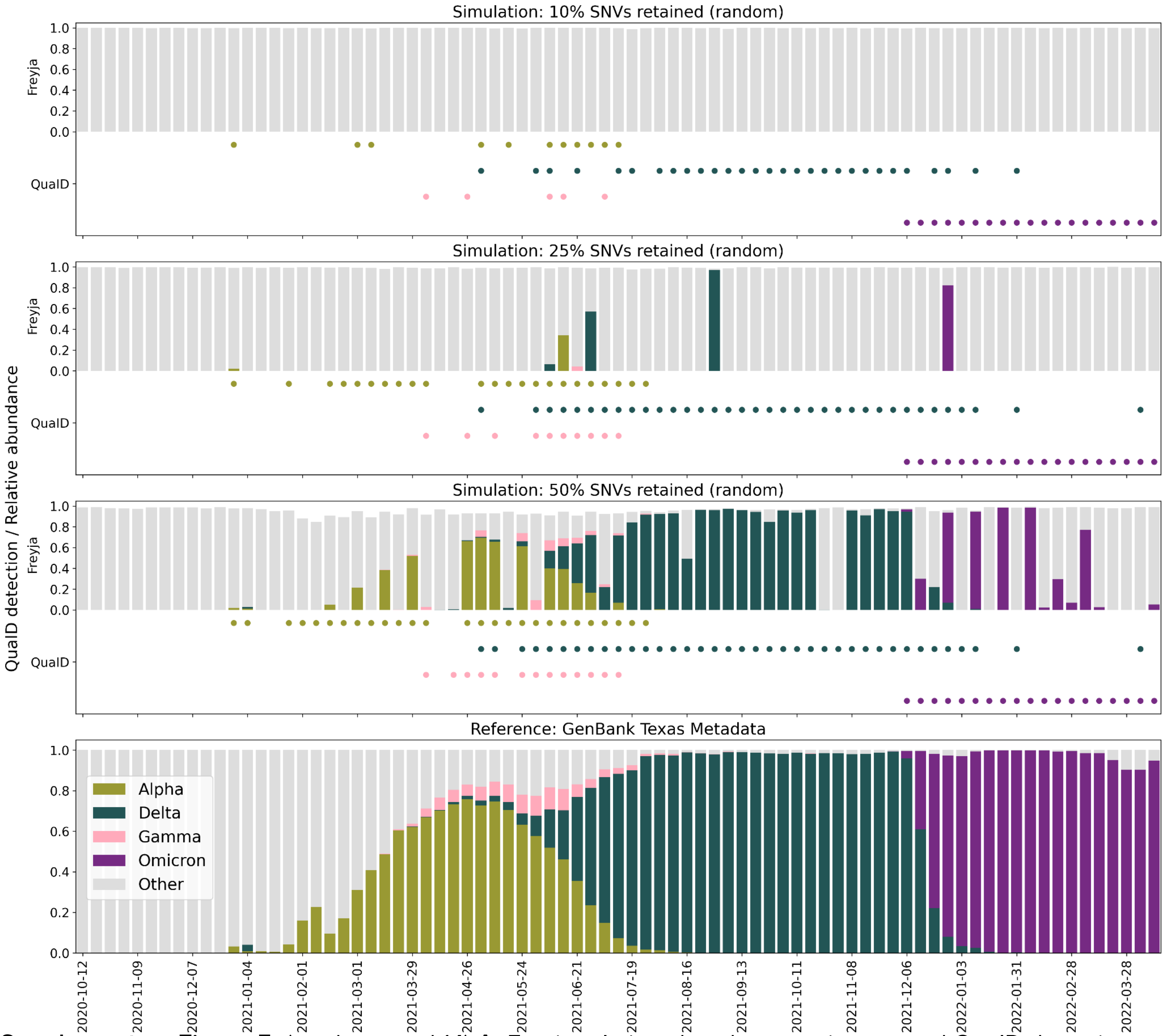


**Supplementary Figure 5.** (random seed 14) **A.** Freyja relative abundance estimates and QuaID detection signal on simulated data from GenBank (USA/TX) with 10% of all SNVs retained at random. Freyja is unable to detect any of the four (Alpha, Delta, Gamma, Omicron) VoCs. QuaID detection is sparse, in particular for the Gamma variant. **B.** Freyja relative abundance estimates and QuaID detection signal on simulated data from GenBank (USA/TX) with 25% of all SNVs retained at random. Freyja sparsely detects major VoCs (Delta, Omicron). QuaID detections become less sparse for all VoCs. **C.** Freyja relative abundance estimates and QuaID detection signal on simulated data from GenBank (USA/TX) with 50% of all SNVs retained at random. Freyja detections become dense, and in some cases abundance estimates correctly reflect simulated abundance profiles. QuaID remains highly sensitive with respect to early detection. **D.** Metadata from GenBank (USA/TX) showing the fraction of genomes belonging to different VoCs for any given week. In this simulated experiment the fractions shown correspond to true relative abundances in the simulated mixture.

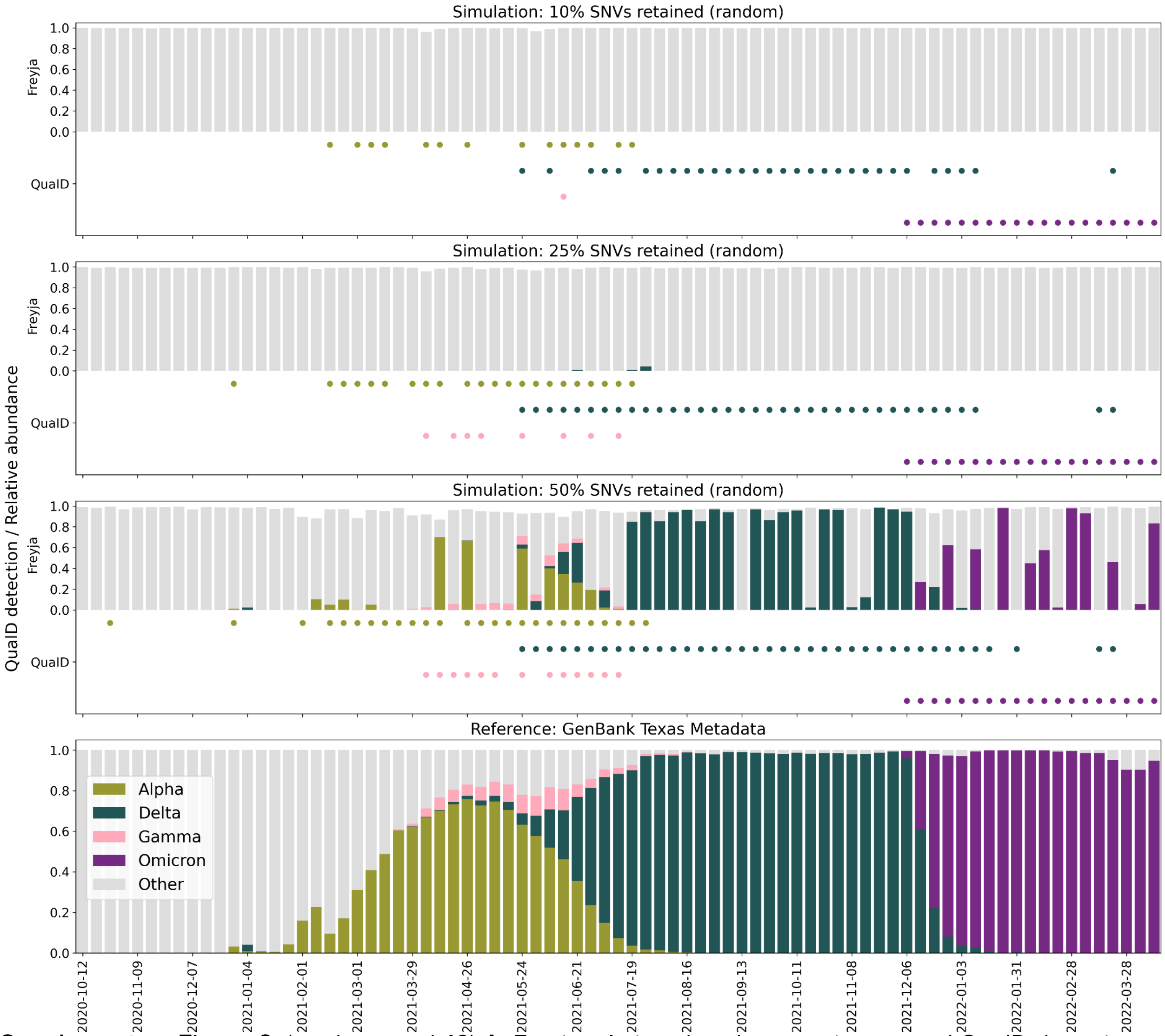


**Supplementary Figure 6.** (random seed 42) **A.** Freyja relative abundance estimates and QualD detection signal on simulated data from GenBank (USA/TX) with 10% of all SNVs retained at random. Freyja is unable to detect any of the four (Alpha, Delta, Gamma, Omicron) VoCs. QualD detection is sparse, in particular for the Gamma variant. **B.** Freyja relative abundance estimates and QualD detection signal on simulated data from GenBank (USA/TX) with 25% of all SNVs retained at random. Freyja sparsely detects major VoCs (Delta, Omicron). QualD detections become less sparse for all VoCs. **C.** Freyja relative abundance estimates and QualD detection signal on simulated data from GenBank (USA/TX) with 50% of all SNVs retained at random. Freyja detections become dense, and in some cases abundance estimates correctly reflect simulated abundance profiles. QualD remains highly sensitive with respect to early detection. **D.** Metadata from GenBank (USA/TX) showing the fraction of genomes belonging to different VoCs for any given week. In this simulated experiment the fractions shown correspond to true relative abundances in the simulated mixture.
